# Direct detection of Dengue viruses without extraction of RNA on the mobile real-time PCR device

**DOI:** 10.1101/2020.11.04.20209635

**Authors:** Masaaki Muraoka, Yukiko Tanoi, Tetsutaro Tada, Aya Tabata, Mikio Mizukoshi, Osamu Kawaguchi

**Affiliations:** Business Innovation Center, Nippon Sheet Glass Co. Ltd., 5-8-1, Nishi-Hashimoto, Midori-ku, Sagamihara-shi, Kanagawa, Japan; Earth Corporation, 12-1, Kanda-Tsukasamachi 2 chome, Chiyoda-ku, Tokyo, Japan; Certified Non-Profit Organization Biomedical Science Association, 2-20-8, Kamiosaki, Shinagawa-ku, Tokyo, Japan

**Keywords:** dengue virus, RNA virus, real-time RT-PCR, direct RT-PCR, rapid, quick, simple, easy, mobile

## Abstract

Dengue virus (DENV) is the cause of dengue / severe dengue and a virus of the Flaviviridae family, furthermore, dengue fever has rapidly spread in the world in recent decades. DENV is transmitted by female mosquitoes, mainly of the specie *Aedes aegypti*. The main method to control or prevent the transmission of DENV is to combat the mosquito vectors. Among these, one of important methods is to monitor the DENVs in the mosquito vectors.

For the detection of DENV, nucleic acid amplification tests (NAAT) were recommended, of which criterion standard is real-time RT-PCR with highly sensitive and specific. However, it takes long time as to judge the result per a reaction, besides the necessity of the treatment of RNA in advance, example of extraction, concentration and purification.

It was our object in this time to develop the method of real-time RT-PCR detecting DENVs in shorter time, moreover without especial treatment of RNA from the mosquito in advance. Besides, this work was performed with combing the mobile real-time PCR device with the one-step RT-PCR reagent.

Firstly, we succeeded in shortening the time of real-time RT-PCR for the detection of DENV per one reaction, so that the judgement needed less than 20 minutes if genomic RNA treated in advance. Moreover, each value on the real-PCR device was quantitatively correlated with the positive control RNA from 1.0 × 10 ^ 3 copies to 1.0 × 10 ^ 0 copies per reaction (This correlation coefficient R^2^ > 0.95). Additionally, it made sure that this method could be applied to each DENV serotype.

Secondly, we established the basis of procedure for the real-time RT-PCR without the treatment in advance so-called “direct”. As the result that the positive control RNA additive was utilized instead of the real DENV, spiked into the mosquito homogenized and sampled the supernatant without treatment, it was possible to detect on the real-time RT-PCR even if mosquitoes immediately after blood-feeding. For this reason, this method might be able to utilize in human sera, too.

According to the results of this work, we could suggest the method is possible to detect DENV more quickly and more simply than heretofore. The Real-time “direct” RT-PCR, especially, could be performed with mobile real-time PCR PCR1100 device and one step RT-PCR reagent only. This method must help to detect some viruses other than DENV, too.

## INTRODUCTION

The World Health Organization (WHO) reported that dengue fever has rapidly spread in the world in recent decades [1]. Dengue virus (DENV) is the cause of dengue / severe dengue and a virus of the Flaviviridae family. Dengue has a wide spectrum of disease, but a vast majority of cases are asymptomatic or mild. Some people develop severe dengue, of which symptom is complications associated with severe bleeding, organ impairment and/or plasma leakage. The pathogenesis of DENV infection is complicated and not well understood. Data suggest that viral, immunopathogenic and other host factors have a role in disease severity. The main risk factors of severe disease include the strain of virus, previous infection with a heterotypic DENV, age and genetic background of the person [2].

DENV is transmitted by female mosquitoes mainly of the specie *Aedes aegypti*, which is also vector of other Flaviviridae family such as Japanese encephalitis, chikungunya, yellow fever and Zika viruses. There are four distinct serotypes of the virus that cause dengue: DENV-1, DENV-2, DENV-3 and DENV-4. Subsequent infections (secondary infection) by other serotypes increase the risk of developing severe dengue [1].

The main factors, of which DENV is rapidly spread in the world, would be rainfall, temperature, urbanization and distribution of the principal mosquito vector *Aedes aegypti* [3]. The other main factors responsible for the geographic rapid spread are also movements of populations or individual via travel [4]. DENVs achieved distribution throughout the tropics and the subtropics region. Moreover, the globalization also enabled introducing multiple viral serotypes co-circulating. These facts might be associated with development of severe dengue for some people.

The WHO makes recommendations that at present, the main method to control or prevent the transmission of dengue virus is to combat the mosquito vectors [1] as follows: (1) Prevention of mosquito breeding; (2) Personal protection from mosquito bites; (3) Community engagement; (4) Reactive vector control; and (5) Active mosquito and virus surveillance. Among these, about (5) is explained as follows: Active monitoring and surveillance of vector abundance and species composition should be carried out to determine effectiveness of control interventions; and Prospectively monitor prevalence of virus in the mosquito population, with active screening of sentinel mosquito collections; These recommendations mean it is important to monitor the DENVs in the mosquitos.

For the detection of DENV, nucleic acid amplification tests (NAAT) were recommended by the WHO [5]. The criterion standard of NAAT for DENVs is real-time RT-PCR, which is highly sensitive and specific, but takes 60–90 minutes to detect DENVs RNA [6-8]. Also several applications of RT-Loop mediated isothermal amplification (RT-LAMP) have been described it possible to detect and differentiate DENV serotypes [9-11], but tests run for more than 30 minutes. Furthermore, real-time RT-recombinase polymerase amplification (RT-RPA) assays take within 10 minutes, but which sensitivities of each of DENV serotypes are not well compared with real-time RT-PCR [12]. Another reported, field-formatted RT-PCR for the dengue virus serotype identification in field-collected mosquitoes under field-deployed conditions too, it takes about 2 hours [13]. Furthermore, any method reported at the above need the treatment of RNA in advance such as concentration, purification and extraction, which must take more time and labour. Previously reported it possible to detect the DENVs in the mosquito by RT-PCR without treatment of RNA in advance, so-called direct RT-PCR [14,15]. After all, it would take more time and labour to judge results by means of not detecting by real-time.

It was our object in this time to develop the method of real-time RT-PCR detecting DENVs for shorter time, moreover without especial treatment of RNA from the mosquito in advance. As follows in this paper, we suggest “quicker and simpler” method, suitable for the field, and useful to effective control in the mosquitoes.

## MATERIALS AND METHODS

### Primer and Probe

To detect all four DENV serotypes by NAAT, we adopted the real-time RT-PCR by hydrolysis probes commonly also referred to as Taqman™, of which commercial kit (LightMix® Modular Dengue Virus, F. Hoffmann-La Roche Ltd., Switzerland) was used. This kit was contained primer / probe mix (a red cap: The Parameter-Specific Reagents), probe of which labelled JOE™ fluorescent dye at 5’ - end. According to references [16] of this kit, NATT was targeted in the 3’untranslated region (UTR) gene sequence unique to a DENV type I∼IV group.

### Sample

Positive control RNA against all four DENV serotypes in the real-time RT-PCR at this works was accommodated in commercial kit of primer / probe kit as mentioned above (a black cap: The Positive Control), of which concentration was adjusted to 1.0 × 10 ^ 3 copies / μL followed by the preparation of 10-fold serial dilution with 10mM Tris-HCl (pH 8.0 adjusted from 1M [Nippon Gene Co., Ltd., Japan] with water [UltraPure™ DNase/RNase-Free Distilled Water, Thermo Fisher Scientific Inc., USA]).

The genomic RNA extracted from each DENV serotypes I, II, III, and IV (unknown about each strain) cultivated were kindly provided by a third party. By commercial kit (QIAamp Viral RNA Mini Kit, QIAGEN GmbH, Germany) was used in the extraction of each, followed by store at -80 °C. Each concentration made a relative evaluation to the above positive control by real-time RT-PCR in the present work. Before assessed by real-time RT-PCR, each was thawed and diluted with 10 mM Tris (pH 8.0) by 100 times.

The mosquito species *A. aegypti* for experiment (pathogen-free bred for many generations at Earth Chemical Co. Ltd., Japan) was used in the present work, and assessed about female within 7 days after adult. More than 10 mosquitoes alive were randomly divided into 3 groups, subsequently only 1 group stored at -80 °C (sample for not blood-feeding). The rest 2 groups were fed blood. About blood-feeding, 3 healthy persons voluntarily participated to be bitten by each mosquito. After each mosquito fed blood a time only, one group was stored at -80 °C immediately after blood-feeding (sample for 0hr after blood-feeding), and another group was cultivated at 28 ± 1 °C for 24 hrs followed by storing at -80 °C (sample for 24 hrs after blood-feeding). Each -80 °C-stored mosquito sample was proceeded at 4 °C as follows: a sample was thawed, transferred into a 1.5 mL microtube, added 99 μL of 10 mM Tris (pH 8.0) and 1μL of 1.0 × 10 ^ 3 copies / μL positive control RNA described above (from LightMix® Modular Dengue Virus kit), homogenized by a disposable homogenizer pestle for 60 seconds and centrifuged briefly. The volume of positive control RNA additive was according to the reference [17], specifically, we judged that DENV titer was at least about 10 ^ 3 PFU / mosquito from the result at kinetics of DENV-2 replication in orally infected mosquitoes. Therefore, the positive control RNA additive was used by 1.0 × 10 ^ 3 copies per a mosquito sample. Subsequently, the upper aqueous were transferred into another 1.5mL microtube and used in each detection by the real-time RT-PCR without extraction of RNA. At the above procedure, 3 mosquito samples were applied per each group.

### Real-time RT-PCR

As previously reported [18], RT and PCR were carrying out in the one-step with using mobile real-time PCR device PCR1100 (Nippon Sheet Glass Co. Ltd., Japan) and one step RT-PCR reagent (THUNDERBIRD® Probe One-step qRT-PCR Kit, TOYOBO Co. Ltd., Japan) for all tests of this study. Briefly describe the composition of RT-PCR reagent, modified as previously reported [18], 1 μL of a sample (positive control RNA, extracted RNA samples or mosquito samples) was amplified in a 16 μL reaction solution containing 1x Reaction Buffer, 1.0 μL of RT Enzyme Mix, 1.0 μL of DNA Polymerase (THUNDERBIRD® Probe One-step qRT-PCR Kit), 0.5 μL of primer / probe mix for targeting all four DENV serotypes (LightMix® Modular Dengue Virus) and 0.3 μM probe (sequence: AGACGCAATACCGCGAGGTG) double-labelled (FAM labelled at 5’end, BHQ® [Black Hole Quencher®, Biosearch Technologies, Inc., USA] labelled at 3’end) for movement of PCR1100. With preliminary tested, each composition was optimized (data not shown).

The RT-PCR condition applied in present work was programmed as follows: RT incubation and enzyme activation were serially performed at 42 °C for 300 seconds, at 95 °C for 15 seconds respectively. Subsequently, thermal cycling was then performed at 95 °C for 3.5 seconds (denaturation), and at 60 °C for 15 seconds (annealing and amplification) for fifty cycles. With preliminary tested, each condition was optimized (data not shown). Incidentally, the procedure of PCR1100 was according to previously reported [19].

## RESULTS

### Detection of DENV in the positive control RNA

Firstly, it needed to determine an evaluation index for properly evaluating each DENV sample by the real-time RT-PCR with the mobile real-time PCR1100 (Nippon Sheet Glass Co. Ltd.) and one step RT-PCR reagent (THUNDERBIRD® Probe One-step qRT-PCR Kit, TOYOBO Co. Ltd.). Therefore, in the present work, that was assessed by using a commercial kit (LightMix® Modular Dengue Virus, F. Hoffmann-La Roche Ltd.). As the result of detecting the positive control RNA accommodated in the above kit by 10-fold serial dilution method, it was possible to detect 1.0 × 10 ^ 0 copies per reaction (fig. 1, Ct = 42.80). Furthermore, each Ct value was quantitatively correlated with the positive control RNA from 1.0 × 10 ^ 3 copies to 1.0 × 10 ^ 0 copies per reaction (the correlation coefficient R^2^ = 0.9844). In this time, the approximate curve was shown as y = -3.998 × + 43.397 (x: log 10 copies of the positive control RNA, y: Ct value, *p* < 0.01), which was used to determine the concentration of RNA for DENV in each sample of the bellow. It took only about 19 minutes per detection with using PCR1100 and one step RT-PCR reagent (THUNDERBIRD® Probe One-step qRT-PCR Kit) in present work.

**Fig. 1.**
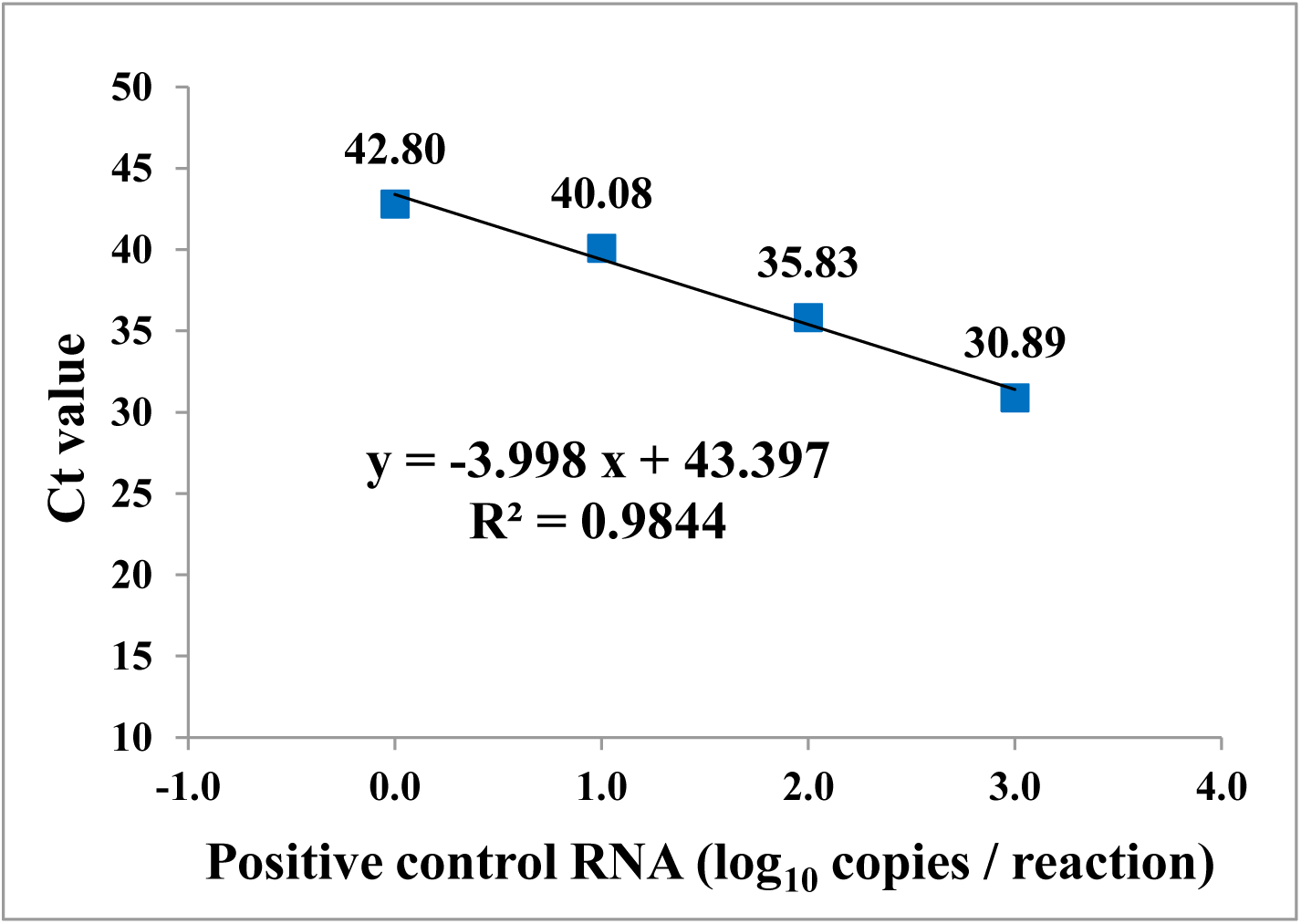
Correlation of copies number and Ct value when detection of positive control RNA on the PCR1100. The approximate curve was shown as y = -3.998 × + 43.397 (x: log10 copies of positive control RNA, y: Ct value, *p* < 0.01), of which relativity was high (this correlation coefficient R^2^ = 0.9844).

### Detection of each serotype DENV-I, -II, -III, and -IV

Each genomic RNA extracted from serotype DENV-I, II, III, and IV was assessed by method as the same of positive control RNA. As the result, it was possible for this method to detect all serotypes (fig. 2). Furthermore, based on the relativity of volume for the positive control RNA to the Ct value (the approximate curve was shown as y = -3.998 × + 43.397 [x: log 10 copies of the positive control RNA, y: Ct value]), the concentration of genomic RNA for each of serotypes was able to be estimated (table 1).

**Table 1.** Results of Ct value for the genomic RNA extracted from each serotype DENV, and each concentration estimated. Based on the relativity of volume for the positive control RNA to the Ct value (the approximate curve was shown as y = -3.998 × + 43.397 [x: log10 copies of the positive control RNA, y: Ct value]), the concentration of genomic RNA for each of serotypes was estimated.

| Serotype | Ct value | Estimated concentration |
| --- | --- | --- |
| DENV-I | 32.8 | 447.3 copies / $\mu\text{L}$ |
| DENV-II | 34.7 | 149.7 copies / $\mu\text{L}$ |
| DENV-III | 33.5 | 298.9 copies / $\mu\text{L}$ |
| DENV-IV | 34.1 | 211.5 copies / $\mu\text{L}$ |

**Fig. 2.**
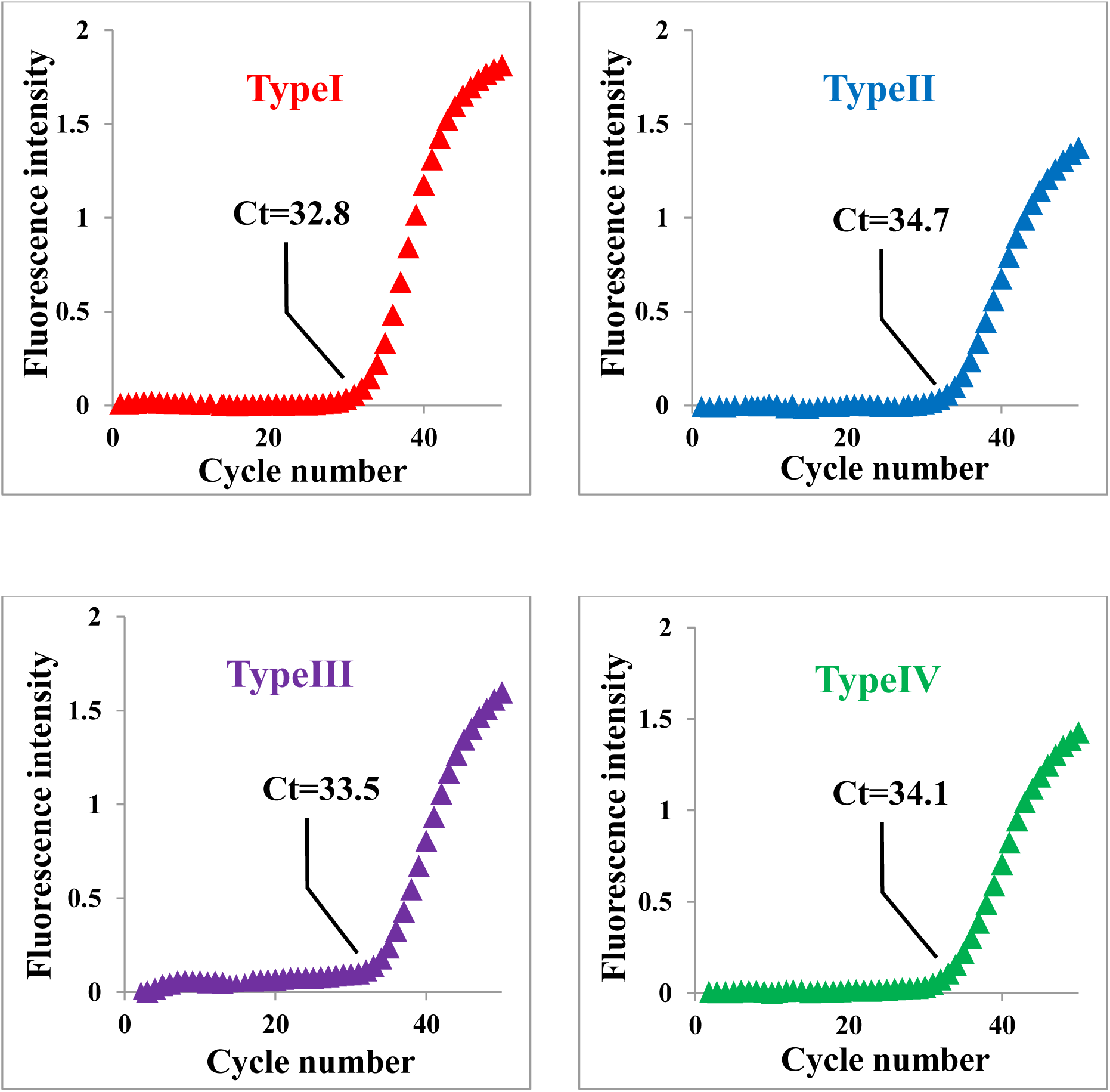
Results of real-time RT-PCR for each genomic RNA extracted from DENV type I: the upper left, type II: the upper right, type III: the lower left, and type IV: the lower right. The Ct value detected was shown on each figure.

### Detection without extraction of RNA from mosquito samples

Using the same method for the real-time RT-PCR above mentioned, the spike method was performed by the positive control RNA additive instead of the real DENVs to assess whether it was possible to detect DENVs by real-time RT-PCR without extraction, concentration and purification of RNA from mosquito samples. Additionally, the mosquito species A. aegypti groups were divided into the group of fed blood or not, which were also assessed it how to influence the real-time direct RT-PCR by blood-feeding. As the result, without treatment for extraction, concentration and purification of RNA, it was possible to detect the positive control RNA in all mosquito samples of group of not fed blood and group immediately after blood-feeding (0 hr) (table 2). There was no significant difference at Ct value between two groups (*p* > 0.05). Furthermore, it was only 1 μL that the volume of each sample was utilized by a reaction of real-time RT-PCR with direct method. On the other hand, the Ct value couldn’t be detected in all mosquito samples of group 24hrs-alive cultivate after blood-feeding. About 2 groups possible to detect Ct value, the real volume of RNA was estimated from each Ct value by the relativity of volume of the positive control RNA to the Ct value (the approximate curve was shown as y = -3.998 × + 43.397 [x: log 10 copies of the positive control RNA, y: Ct value]) (table 2). As the result of each, both samples were shown only less than 20 % compared to the volume of positive control RNA additive for the spike. (fig. 3)

**Table 2.** Results of Ct value for the detection of DENV in each of mosquito samples spiked positive control RNA. *Estimated RNA volume; The volume of RNA possible to be detected in each sample was estimated by the relativity of volume for the positive control RNA to the Ct value (the approximate curve was shown as y = -3.998 × + 43.397 [x: log10 copies of the positive control RNA, y: Ct value])

| Mosquito group | Sample ID | Ct value | *Estimated RNA volume | Estimated RNA mean | Estimated RNA SD |
| --- | --- | --- | --- | --- | --- |
| Not blood-feeding | #1 | 42.36 | 1.82 copies | 1.56 copies | 0.55 |
|  | #2 | 43.51 | 0.94 copies |  |  |
|  | #3 | 42.25 | 1.94 copies |  |  |
| 0 hr after blood-feeding | #4 | 41.48 | 3.02 copies | 2.01 copies | 1.03 |
|  | #5 | 43.47 | 0.96 copies |  |  |
|  | #6 | 42.15 | 2.05 copies |  |  |
| 24 hrs after blood-feeding | #7 | Not detected | Not detected | Not detected | Not detected |
|  | #8 | Not detected | Not detected |  |  |
|  | #9 | Not detected | Not detected |  |  |

**Fig. 3.**
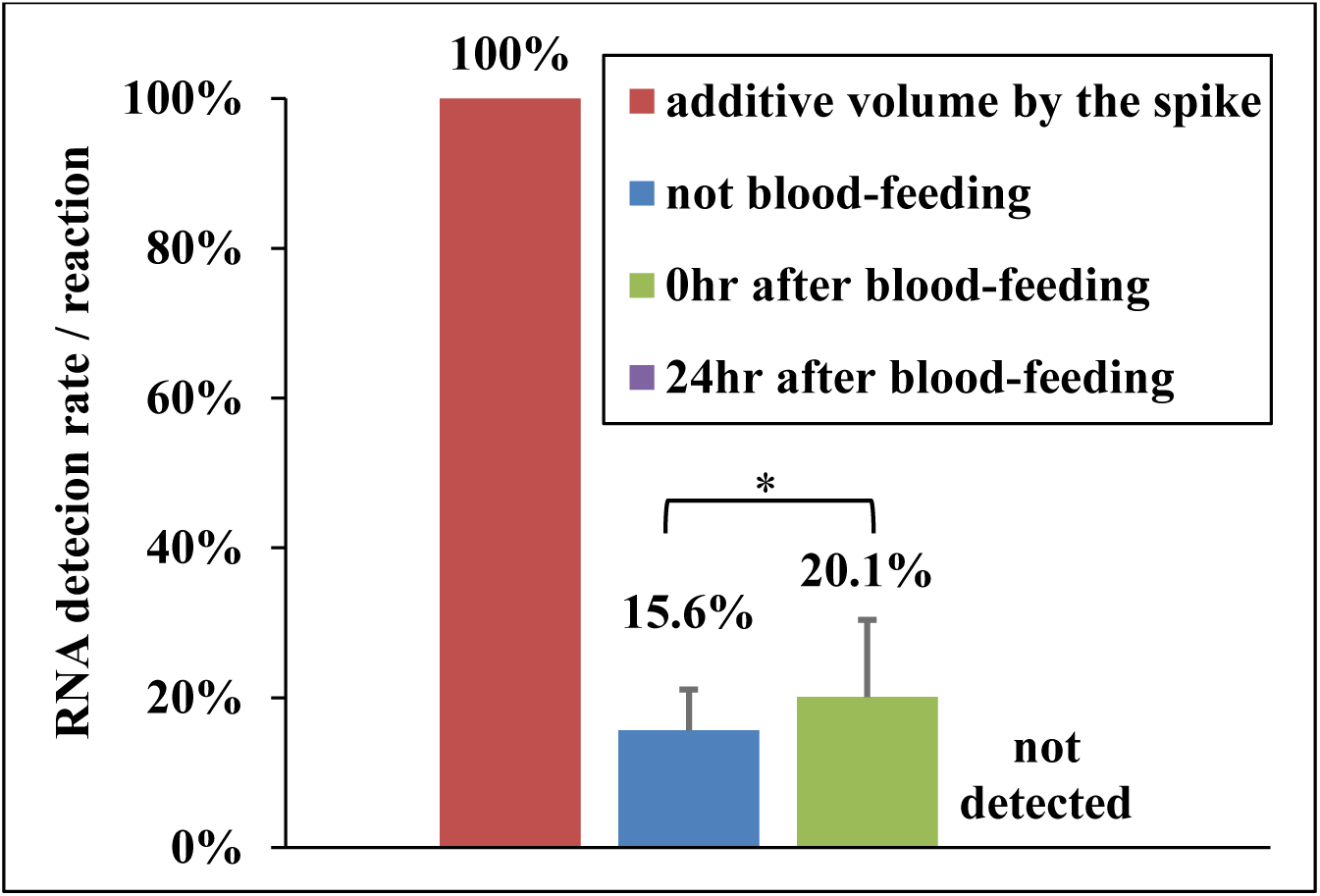
RNA detection rate for dengue virus in the real time RT-PCR of positive control RNA. The additive volume of positive control RNA for DENV by the spike was correspond to 10 copies per a RT-PCR according to the above methods. Based on the table 2, for each of groups, fig.3 showed the relative detection in each group taking the level in additive volume of positive control RNA as a base of 100. *; There was no significant difference between two groups (*p* > 0.05)

## DISCUSSION

In this work, we had main objects to construct new methods for detection of DENVs by real-time RT-PCR without extraction, concentration and purification of RNA from each mosquito sample, and to assess whether it was possible to quickly and simply judge results. Therefore, referenced the method as previously reported [18], this time such the same, it utilized to be combined the mobile real-time PCR device PCR1100 (Nippon Sheet Glass Co. Ltd.) with one step RT-PCR reagent (THUNDERBIRD Probe One-step qRT-PCR Kit [TOYOBO Co. Ltd.]) this time too.

Firstly, the method reported previously was modified a part to be suitable for the commercial kit for detection of DENVs. LightMix® Modular Dengue Virus (F. Hoffmann-La Roche Ltd.) was selected as the commercial kit for detection of DENVs this time, because of certification for detection at the relativity of volume for the positive control RNA on another real-time PCR device LightCycler® (F. Hoffmann-La Roche Ltd., Switzerland). As this result, also on the PCR1100 device, it was proved that the kit selected was able to be utilized, furthermore with high relativity (This correlation coefficient R^2^ > 0.95) detected till at least 1.0 × 10 ^ 0 copies of the positive control RNA for DENV the same as on another device. Accordingly, we determined to able to utilize this method to assess subsequent correctly.

At next stage, by methods as mentioned above, it was evaluated whether it was possible to detect the genomic RNA for each of DENV serotypes in practice. As the result, in all DENV serotypes, it became clear that it is possible to detect from the genomic RNA by this method for real-time RT-PCR. According to the fist result of positive control RNA, the concentration of genomic RNA for each serotype could be calculated. In this time, because each sample was known for the serotype, it was not necessary to corroborate each type. However, if there is no information about what kind of serotype, the PCR1100 device might quickly and simply discriminate serotype by multi-channel detection system for example the methods in Centers for Disease Control and Prevention in USA (CDC) [8]. This theme should be assessed hereafter, of which methods would be efficient to monitor DENV serotypes and to suppress severe disease including the strain of virus and previous infection with a heterotypic DENVs [2].

Additionally, it took only less than 19 minutes to finish out in this method, which was very quick compared with other current methods for real-time RT-PCR (for 70 minutes if using LightCycler® according to method of LightMix® Modular Dengue Virus). If already finished extracting the genomic RNA from each sample, this device would be quickly able to discriminate the presence of target. This faster RT-PCR might be caused by combination of PCR1100 with the one-step RT-PCR reagent (THUNDERBIRD® Probe One-step qRT-PCR Kit), therefore it was efficiently possible to carry out RT and PCR at one step in sequence. In addition, if each condition of RT-PCR, especially time of annealing and amplification could be validated, total time of a real-time RT-PCR might be shortened.

In the next try, we assessed the method for real-time RT-PCR without treatment of RNA in advance, so-called real-time direct RT-PCR. The spike method was performed by using the positive RNA additive instead of the real DENVs this time. As the result, in the mosquitoes immediately after blood-feeding, it was possible to adequately detect the positive control RNA additive without treatment of RNA in advance, but a fifth of additive only could be detected calculatedly. Because it was similar in the mosquitoes not fed blood, each real-time RT-PCR might be inhibited by something in the mosquito, example of the nuclease, the proteinase, the absorption and the pH changer. Moreover, in all mosquito samples of group 24hrs-alive cultivate after blood-feeding, Ct values were not detectable. It might be the cause that the inhibitors described above increased in individual mosquito after blood-feeding. Actually, it has been reported that the blood induced various enzyme in the mosquitoes several hours after biting [20]. The virus to actual transmission to a new host is termed the extrinsic incubation period (EIP), which the EIP for DENVs takes about 8-12 days when the ambient temperature is between 25-28 °C [1]. With these taken into consideration, we must assess about different times of alive cultivate after blood-feeding.

In the first try, this time, the volume of positive control RNA additive was according to the reference [17]. Thus, the positive control RNA additive was used by 1.0 × 10 ^ 3 copies per a mosquito sample. As the result, it was possible to detect about 2.0 copies (more than 1.0 copies, more less 3.0 copies) per a reaction by this method. Accordingly, at this time only if used the mosquitoes immediately after blood-feeding as the sample, this method could be used as the real-time “direct” RT-PCR. As described above, when the EIP for DENVs takes about 8-12 days [1], the DENV titer also amplify about 10 ^ 4 PFU / mosquito [17]. Therefore, we would have to assess spike test with this consideration too.

Additionally, by focusing on that this method can detect even if much human blood fed to a mosquito, it might be utilized at human blood. It has been reported that viral RNA detected were 1.04 × 10^4^ ∼ 6.9 × 10^12^ copies / mL in sera from dengue patients for DENV-1 to 4 during 2–5 days post onset of fever [21], while the 50% mosquito infectious dose for each of DENV-1-4 ranged from 6.29 to 7.52 log10 RNA copies/mL of plasma [22]. Taking concentration of virus in sera into consideration, this method might be applied to detect DENV in human sera as real-time direct RT-PCR quicker and simpler than heretofore.

In this paper, we suggest the methods possible to detect DENV more quickly and more simply compared to previously. Real-time “direct” RT-PCR, especially, could be performed with mobile real-time PCR PCR1100 device and one step RT-PCR reagent only. To ensure this method, we would like to assess about the mosquitoes and the larvae collected in the field, and then about human sera too. At such time, it is very important that these methods should be gotten some experiences to use quickly and simply on site by utilizing the characteristics of mobile real-time PCR device. Incidentally, DENV is the same flavivirus as Zika and chikungunya, and three mosquito-borne viruses are having overlapping transmission vector [23]. For this reason, it is the research task from now on that new methods must be designed with utilizing multi-channel, which could detect three different kinds of viruses simultaneously and simply. These methods should be useful for virological surveillance of Aedes mosquitoes infected something viruses, as an early warning and sustainable system to predict outbreaks of viruses.

## STUDY DESIGN

When each sample in this work was assessed in the real-time RT-PCR (PCR1100, Nippon Sheet Glass Co. Ltd., Japan), a ten-fold dilution series of positive control RNA (LightMix® Modular Dengue Virus, F. Hoffmann-La Roche Ltd., Switzerland) was indicated with high relativity to Ct value on the mobile real-time PCR device PCR1100 (This correlation coefficient R^2^ > 0.95) the same as on the another device.

## Data Availability

The data that support the findings of this study are available from the corresponding author upon reasonable request.

## ACKNOWLEDGEMENTS

We thank volunteers participated to be bitten by mosquitoes, Shunsuke Sejima at Certified Non-Profit Organization Biomedical Science Association for technical comments, and Takashi Fukuzawa in an ex-employee at Nippon Sheet Glass Co. Ltd. for technical comments.

## AUTHOR CONTRIBUTIONS

M. Muraoka conceived, designed experiments, performed experiments, analysed the data and wrote the manuscript. Y. Tanoi performed experiments and analysed the date. T. Tada wrote the manuscript. A. Tabata conceived and provided specimens. M. Mizukoshi conceived. O. Kawaguchi conceived, designed experiments. All authors also participated in the editorial process and approved the manuscript.

## ETHICS STATEMENT

The study was approved by the Ethics Committee of Certified Non-Profit Organization Biomedical Science Association (BMSA2020-11-01). Written informed consent was obtained from each volunteer participated to be bitten by mosquitoes.

